## Supplementary materials for "Cardiovascular vulnerability predicts hospitalisation in primary care clinically suspected and confirmed COVID-19 patients: a model development and validation study"

**Supplementary table 1** – ICPC codes used in this study

| **Inclusion** | **ICPC code** |
| --- | --- |
| Suspected COVID-19 | R81, R81.00, R81.01 |
| Confirmed COVID-19 | R83, R83.00, R83.01, R83.02, R83.03 |
| Acute upper respiratory infection | R74, R74.00, R74.01, R74.02 |
| **Comorbidities** | **ICPC code** |
| History of any cancer* | A79, B72, B72.01, B72.02, B73, B74, B74.01, D74, D75, D76, D77, D77.01, D77.02, D77.03, D77.04, F74.01, H75.01, K72.01, L71.01, N74, R84, R85, S77.03, S77.04, T71, U75, U76, U77, W72, X75, X76, X76.01, X77, X77.01, X77.02 |
| Hypercholesterolemia | T93, T93.01, T93.02, T93.03, T93.04 |
| Heart failure | K77, K77.01, K77.02 |
| Hypertension | K85, K86, K87 |
| Ischaemic heart disease | K74, K74.01, K74.02, K75, K76, K76.01, K76.02 |
| Type 2 diabetes mellitus | T90, T90.02 |
| Peripheral arterial disease | K91, K92 |
| History of stroke/TIA | K89, K90, K90.03 |
| History of VTE | K93, K94, K94.01, K94.02, W77.03, W99.03 |
| Atrial fibrillation | K78 |
| COPD | R91, R91.01, R91.02, R95 |
| Asthma | R96, R96.02 |

*excluding skin cancers except for melanoma

ICPC codes used for identifying the study population and individual comorbidities.
COVID-19 = coronavirus disease 2019; ICPC = International Classification of Primary Care; TIA = transient ischemic attack; VTE=venous thromboembolism.

**Supplementary figure 1** – Calibration plots in individual databases

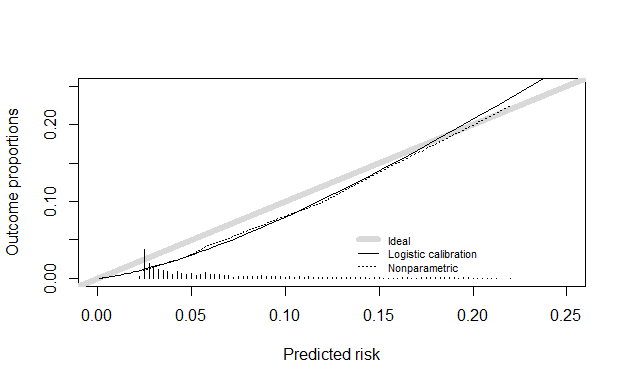

Supplementary figure 1a - Calibration plot in the total validation cohort with hospitalisation as the outcome

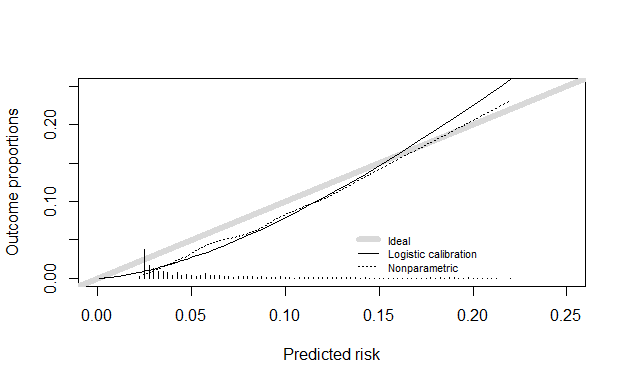

Supplementary figure 1b – Calibration plot in JGPN validation cohort with hospitalisation as the outcome

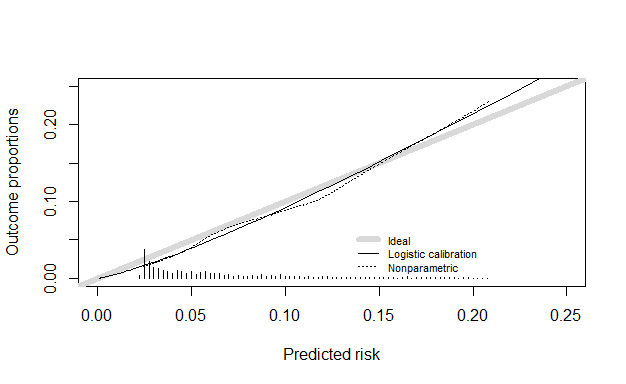

Supplementary figure 1c – Calibration plot in AHA validation cohort with hospitalisation as the outcome

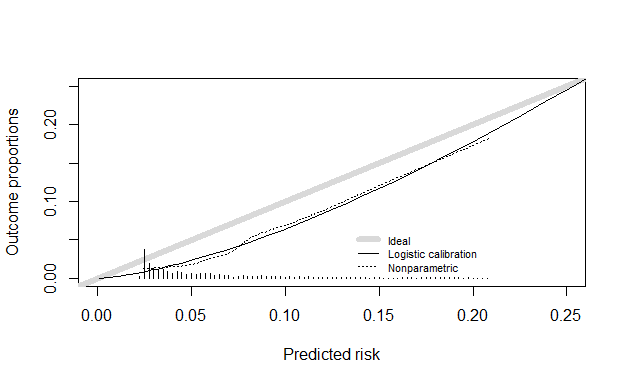

Supplementary figure 1d – Calibration plot in ANH validation cohort with hospitalisation as the outcome
